# Have news reports on suicide and attempted suicide during the COVID-19 pandemic adhered to guidance on safer reporting? A UK-wide content analysis study

**DOI:** 10.1101/2021.04.19.21255736

**Authors:** L. Marzano, M. Hawley, L. Fraser, E. Harris-Skillman, Y.X. Lainez, K. Hawton

**Affiliations:** Faculty of Science and Technology, Middlesex University, The Burroughs, Hendon, London NW4 4BT; Samaritans, The Upper Mill, Kingston Road, Ewell, Surrey, KT17 2AF; Centre for Suicide Research, Department of Psychiatry, University of Oxford, Warneford Hospital, Oxford, OX3 7JX; Oxford Health NHS Foundation Trust, Warneford Hospital, Oxford, OX3 7JX

**Keywords:** suicide, media, newspaper, reporting, media guidelines, COVID-19

## Abstract

Associations between sensational news coverage of suicide and subsequent increases in suicidal behaviour in the general population have been well documented. Amidst growing concern over the impact of the COVID-19 pandemic on suicide rates, it is especially important that news coverage of suicidal behaviour adheres to recommended standards for the responsible reporting of suicide. Using a set of dimensions based on international media guidelines, we analysed the quality and content of all UK news reports of possible COVID-19 related suicides and suicide attempts in the first four months of the pandemic (N=285 reports of 78 individual incidents published in print and online newspapers between 16^th^ March and 12^th^ July 2020). The majority of news reports made an explicit link between suicidal behaviour and the COVID-19 pandemic in the headline (187/285, 65.5%), and portrayed this association as strong and direct (n=196/272, 72.1%), mostly based on statements by family, friends or acquaintances of the deceased (171/285, 60%). The impact of the pandemic on suicidal behaviour was most often attributed to feelings of isolation (78/285, 27.4%), poor mental health (42, 14.7%) and sense of entrapment (41, 14.4%) as a result of government-imposed restrictions. Although rarely of poor overall quality, reporting was biased towards young people, frontline staff and relatively unusual suicides (including those involving a celebrity, murder-suicide and violent methods) Also, to varying degrees, reports failed to meet recommended standards; for example, 41.1% (117/285) did not signpost readers to sources of support, a quarter (69, 24.2%) included examples of sensational language and a third provided over-simplistic explanations for the suicidal behavior (93, 32.6%). While news reporting has improved compared to earlier coverage of suicide in the UK, it is essential that careful attention is paid to the quality and content of reports, especially as longer-term consequences of the COVID-19 pandemic develop.

## 1. Introduction

News reporting of suicidal behaviour can have important influences on population levels of suicide and self-harm (Sisask and Värnik, 2012). This especially applies to reports which are dramatic, extensive, on the front page of newspapers, and include a sensational headline, which can lead to increases in suicide (Niederkrotenthaler et al., 2010; Pirkis et al., 2006). Reports of suicides of celebrities appear to be particularly powerful in influencing suicide in the general population, especially when the method of suicide is highlighted (Niederkrotenthaler et al., 2020). The highly publicised deaths of the actor Robin Williams (Fink et al., 2018) and of the German national football team goalkeeper Robert Enke (Koburger et al., 2015) are examples. News reporting of suicide is disproportionately focussed on deaths of young people and on females relative to their actual involvement in suicide (Marzano et al., 2018). Young people are also most susceptible to the influence of news reports on suicidal behaviour (Gould et al., 2014). This is particularly concerning as suicide rates in young people have increased substantially in recent years, at least in the UK (Bould et al., 2019; Office for National Statistics, 2020).

Times of crisis that might have an influence on suicide rates are periods when the impact of news reporting on suicide may be especially important. Concerns have been expressed that the COVID-19 pandemic may cause a rise in rates of both suicide and self-harm (Gunnell et al., 2020; Reger et al., 2020). Fortunately, until at least late summer 2020, this had not occurred in most countries with real-time suicide data (John et al., 2020; Leske et al., 2021; Qin and Mehlum, 2021), although in Japan there was an increase in suicides in females and young people in the latter part of 2020 (Tanaka and Okamoto, 2021). On top of the large numbers of deaths due to COVID-19 infection and the social impacts of the pandemic, such as social isolation and loneliness, the major longer-term impacts of the pandemic on employment, finances and population morale may mean that a future increase in suicide and attempted suicide is more likely, especially in view of the impact of the recent great recession and previous recessions on suicidal behaviour (Gunnell and Chang, 2016; Hawton et al., 2016; Oyesanya et al., 2015). This means that responsible news reporting of suicides is even more of a priority during this time (Hawton et al., 2021).

Because of increasing recognition of the importance of news reporting of suicide in relation to population risk of suicidal behaviour, encouragement of responsible media reporting is included as an important issue in national suicide prevention policies (Department of Health and Social Care, 2012). There are also well-established guidelines for media reporting on suicidal behaviour (Samaritans, 2020a; World Health Organization (WHO), 2017), together with specific guidance about such reporting in relation to the COVID-19 pandemic (Hawton et al., 2021; Reidenberg and Niederkrotenthaler, 2020).

We have carried out a study of news reporting of suicide and attempted suicide in the UK during the first 4 months of the COVID-19 pandemic. News reports of individual suicides in both national and local news outlets were identified through a well-established media monitoring service (Fraser et al., 2017). The reports were subjected to systematic data extraction, which reflected recommended standards in media reporting on suicide (Samaritans, 2020; WHO, 2017) and specific concerns about such reports in relation to the pandemic. The overall aim was to determine the extent to which news reporting adhered to recommended standards for reporting of suicides in general and to identify whether there were specific aspects of reporting related to the pandemic which are of particular concern.

## 2. Method

### 2.1 News database and coding

Print and online newspaper reports of fatal and non-fatal suicidal behaviour and suicide inquests in national and regional British publications (circa 6,000-7,000 per year) are monitored on an ongoing basis by the suicide prevention charity Samaritans (Fraser et al, 2017). Electronic press clippings provided by a specialised news monitoring service are coded by trained staff for content, including identification of the article itself (e.g. date, headline and newspaper) and details of the incident/s being reported (e.g. in relation to method, location, and the age and gender of the individuals involved). Based on adherence to media guidelines (e.g. whether the article includes detailed description of the method used, and details of support services and organisations), each news report is also rated for quality (as ‘positive’, ‘neutral’ or ‘negative’) in relation to its headline, imagery (where applicable) and overall tone (see Marzano et al., 2018 for further details).

#### 2.1.1 COVID-19 related news stories

As of March 16^th^ 2020, all articles in the Samaritans’ media monitoring database which include explicit statements or speculation about COVID-19 related influences on suicidal behaviour by individuals have also been coded to identify: 1) the source and nature of such evidence or speculation; 2) which element or elements of the pandemic (and associated challenges and restrictions) are highlighted in headlines, content and images in news stories as having contributed to suicidal behaviour; and 3) which of these (if any) are identified as the ‘main issue’ in relation to specific acts of suicide or attempted suicide. In addition, using a set of dimensions based on international recommendations for responsible reporting of suicide in the COVID-19 pandemic and its aftermath (Reidenberg and Niederkrotenthaler, 2020; Samaritans, 2020b), the database also includes: 4) any specific issues or concerns in relation to individual news stories (e.g. the use of sensational language, simplistic explanations of suicide); and 5) any positive messages being communicated in such reports (e.g. reaching out to loved ones, encouraging help-seeking) (see online appendix for full details of the coding frame).

To ensure that important aspects of the pandemic and its potential influence on suicidal behaviour have been appropriately and comprehensively recorded, the coding scheme was adapted from the ‘Classification of COVID-19 related factors involved in self-harm’ used in an investigation of hospital attendances for self-harm in the Multicentre Study of Self-harm in England (Hawton et al., 2020). For other sections of the scheme, codes (where not binary yes/no categories) were derived inductively for content (Elo and Kyngäs, 2008) and refined through an iterative process of development and piloting (by three independent raters (MH, EHS and YL)), at different stages of the data collection process. Regular team meetings served to reconcile any discrepancies and reach consensus on the final coding scheme.

### 2.2 Data analysis

We analysed the nature and content of all COVID-19 related articles recorded in the Samaritans media monitoring database from the week before the first UK national lockdown came into force (on 23^rd^ March 2020) and the subsequent four months (i.e., all entries between 16^th^ March and 12^th^ July 2020). All data are presented as frequencies or percentages (e.g. of items that deviated or not from recommended standards in relation to specific aspects of reporting of suicide). For some analyses, these are presented both in relation to the individual events reported in the news during this period (hereafter referred to as ‘individual suicides or stories’), and multiple reports of the same incident (hereafter referred to as ‘all reports’).

## 3. Results

Between 16^th^ March and 12^th^ July 2020, there were 285 reports in online and print news of possible COVID-19 related suicidal behaviour, which constituted 21% of all suicide news recorded in the Samaritans’ media monitoring database for this period (N=1,338). Although there were no COVID-19 related reports in the first nine days of monitoring, most days thereafter saw the publication of at least one COVID-19 related story (with the exception of three days in April, ten in May, twelve in June and two in the first half of July), with a peak of 19 COVID-19 related news items on 25^th^ March (two days into the first UK lockdown) and a median of two reports per day over the four month study period (Figure 1). Most articles appeared in tabloid (175, 61.4%) or regional news outlets (69, 24.2%), with far fewer in broadsheets (24, 8.4%) and other media, e.g. consumer magazines (17, 6.0%). The majority were in online news reports (202, 70.9%). Nearly one in ten print articles were located in prominent positions within the paper (in 9/83 print articles these were on pages 1 to 3).

**Figure 1.**
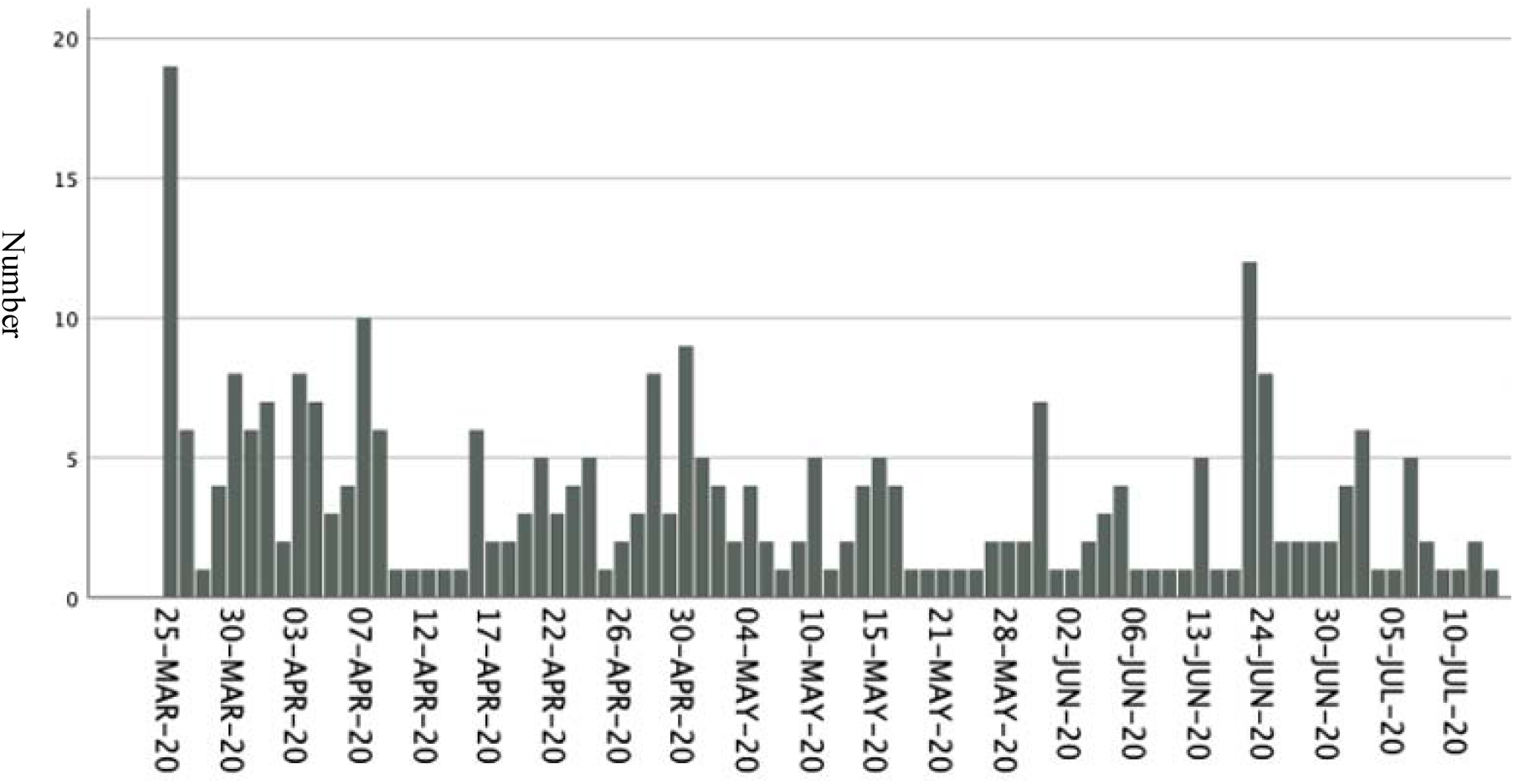
**Daily coverage of possible COVID-19 related suicidal behaviour in national and regional British news (16th March to 12th July 2020)**

### 3.1 Individual suicide stories and repeat reporting

During the study period, 78 individual stories of suicidal behaviour related to COVID-19 were reported in the news. Ninety percent of these were individual suicides (n=65) or attempts (n=6), six were murder suicides and one an alleged suicide pact involving an elderly couple. Most stories were reported around the time of the incident (vs. reports of inquests of earlier deaths or other related events (e.g. funerals, tributes)) (Table 1).

**Table 1.** **News reporting of possible COVID-19 related suicidal behaviour in individual stories and all reports (16**^**th**^ **March to 12**^**th**^ **July 2020): article content and suicide location\***

| Article content | Individual stories<br>(N=78) |  | All reports<br>(N=285) |  |
| --- | --- | --- | --- | --- |
|  | n | (%) | n | (%) |
| Individual suicide/attempt | 71 | (91.0) | 253 | (88.8) |
| 2+ suicides/attempts | 1 | (1.3) | 1 | (0.4) |
| Murder suicide | 6 | (7.7) | 31 | (10.9) |
| Attempted suicide (vs. completed suicide) | 6 | (7.7) | 6 | (2.1) |
| Report type |  |  |  |  |
| Incident | 51 | (65.4) | 202 | (70.9) |
| Inquest | 11 | (14.1) | 26 | (9.1) |
| Other | 16 | (20.0) | 57 | (20.0) |
| Male suicide (vs. female suicide) | 50/76 | (65.8) | 191/282 | (67.7) |
| Suicide location |  |  |  |  |
| England | 30/73 | (41.1) | 137/278 | (49.3) |
| Wales | 8/73 | (11.0) | 47/278 | (16.9) |
| Scotland | 4/73 | (5.5) | 5/278 | (1.8) |
| Northern Ireland | 3/73 | (4.1) | 4/278 | (1.4) |
| USA | 14/73 | (19.2) | 50/278 | (18.0) |
| EU countries | 6/73 | (8.3) | 26/278 | (9.4) |
| Non-EU countries (excluding USA) | 8/73 | (11.0) | 9/278 | (3.2) |

\*Some denominators vary because of information being omitted from news reports.

The vast majority of stories (240/285, 84.2%) appeared in at least 2 news reports (median number of reports per story=2), and 19 were the subject of 5 or more news items (max=17). Murder suicides were somewhat more likely to appear in multiple reports (31 reports of 6 individual stories), as were stories involving (para)medical professionals (46 reports of 9 individual stories), celebrities (31 news items of 4 suicides), and pupils/students (25 reports of 5 individual stories).

COVID-19 related suicide coverage (and repeat coverage) was skewed towards younger age groups (Figure 2), with a small percentage of individual stories and all reports focusing on individuals over the age of 60 years. Approximately two-thirds of individual stories, and reports overall, were about suicidal behaviour amongst men or boys (Table 1).

**Figure 2.**
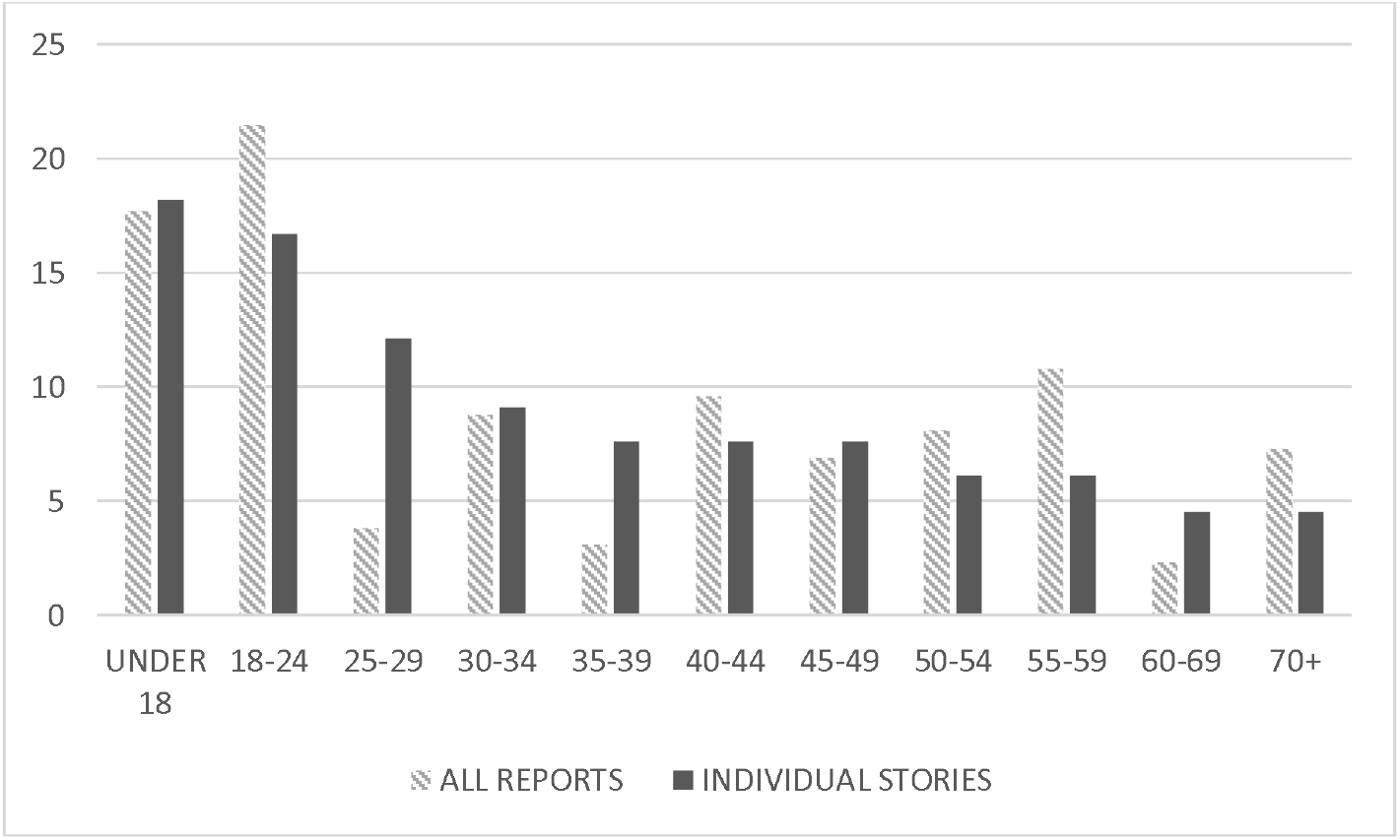
**Percentage of possible COVID-19 related suicide news items focusing on different age groups (where known) in all reports (N=260) and individual stories (N=66)**.

Almost a third of reports focused on suicides and attempts outside the UK (85/285, 29.8%). US-based suicides featured particularly prominently in the British press (18% of all news items were reports of 14 US-based events, including a murder suicide (4 articles) and the death of a celebrity (17 COVID-19 related news items)).

Details of methods used in the suicidal behaviour were included in 105 (37.0%) reports, and in approximately one in ten headlines (26, 9.1%). The most common method of suicide that was reported was jumping/falling from a high place (29, 27% of all reports, and 14/37, 37.8% of individual stories for which suicide method was reported). Deaths by firearms were the second most commonly reported method (22/105 (21.0%) news stories reported 6 deaths by shooting, all but 2 in the US). Suicidal behaviour involving hanging (17 reports of 9 individual stories), self-poisoning (12 reports of 4 individual stories) and self-cutting (8 reports of 3 individual stories) was reported less frequently. There were a few reports of suicides involving less common methods (e.g. self-immolation and drowning).

### 3.2 COVID-19 and suicidal behaviour

Most headlines (187/285, 65.5%) made an explicit link between suicidal behaviour and the COVID-19 pandemic. The terms most frequently used to indicate a possible link were ‘Coronavirus’ (n=96), ‘Lockdown’ (n=95) and ‘virus’ (n=82). ‘COVID’ was infrequently included (n=9). One in five articles (59/285, 20.7%) included one or more COVID-19 related images. These included depiction of current infection rates (n=26), of medical staff or equipment (n=30), or of the virus itself (n=4). Images of the individuals whose suicidal behaviour was reported in the stories featured in 176 (61.8%) articles, and a further 33 (11.6%) articles included images of friends and family of the deceased (particularly in relation to a death of a celebrity). Photos or ‘photo galleries’ of others who died by suicide in similar circumstances appeared in 3 news items (1.1%), whilst images of suicide methods (5, 1.8%), locations (38, 13.3%) and/or other significant places (e.g. the person’s home or workplace (5, 1.8%)) appeared in a total of 43 articles (15.1%).

In the articles themselves, the link between COVID-19 and suicidal behaviour was mostly portrayed as strong and direct (n=196/272, 72.1%). In most cases, the stated link or links between COVID-19 and suicidal behaviour were based on statements made by family and friends of the deceased (157/285, 55.1%), with some links being speculation by neighbours and other acquaintances (n=14). Official sources such as the Police or Coroners were the main source in relation to 33 reports (12.1%), whilst nine others were based on written messages or social media posts by the individual themselves. Press speculation accounted for a fifth of all suggested links (56, 19.6%).

The impact of the pandemic on suicidal behaviour was most often attributed in the news to feelings of isolation (Table 2). A substantial proportion of articles also mentioned other negative outcomes associated with COVID-19, including infection fears, feelings of entrapment and the stress of working on the frontline at this challenging time. Worsening mental health and emerging, pandemic-related mental health issues were also common themes in news reporting of possible COVID-19 related suicides. One in five individuals in such reports was said to have had a history of mental health difficulties (57/285, 20.0%), in some cases in addition to neurodevelopmental (n=7) or substance abuse issues (n=5), trauma (including bereavement) (n=5), or a known history of self-harm (n=5). For some, reduced contact with mental health services as a result of the pandemic was reported as a significant factor in their suicidal behaviour (n=12).

**Table 2.**
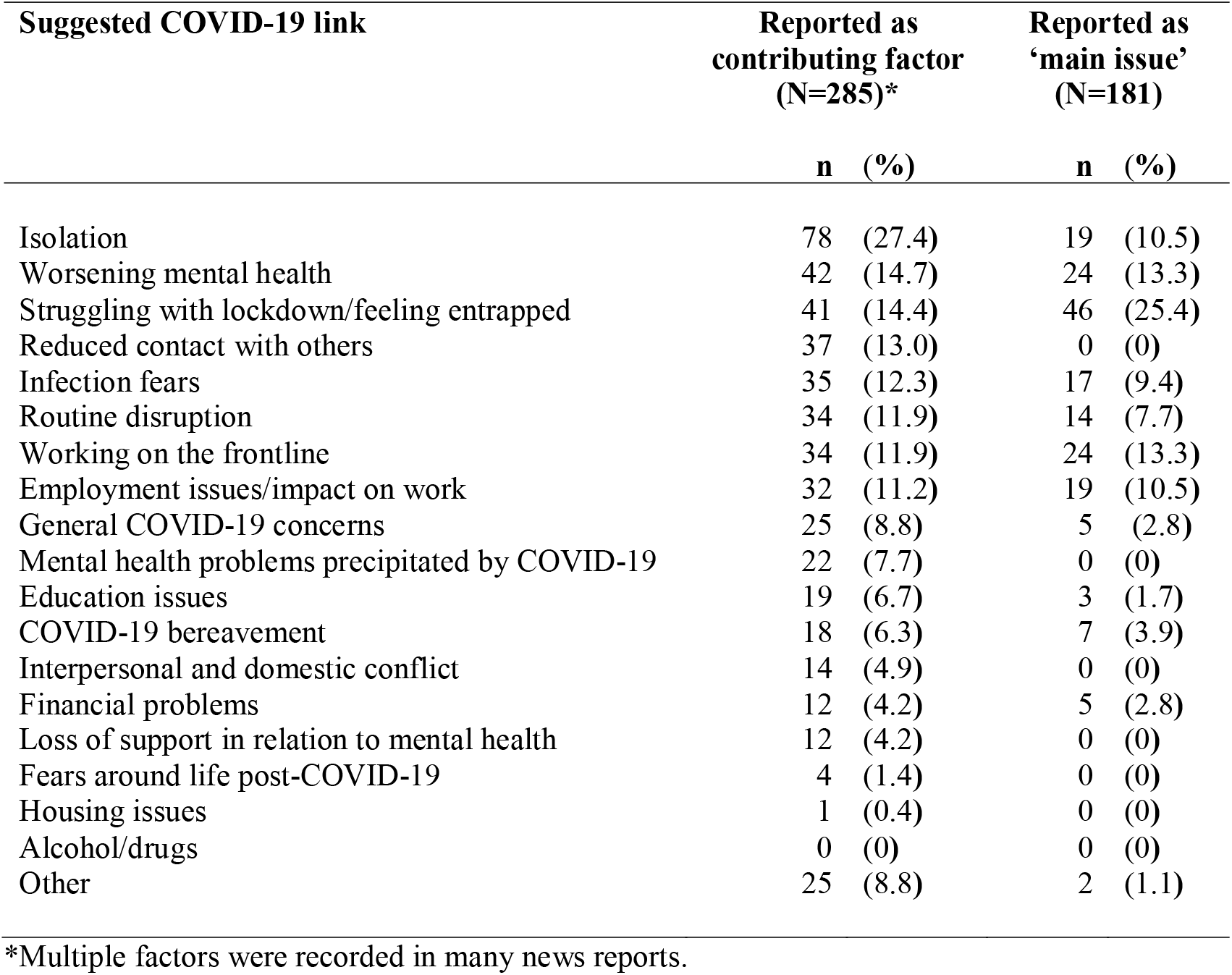
**Suggested COVID-19 related influences on suicidal behaviour in British news reports (16**^**th**^ **March to 12**^**th**^ **July 2020)**

The impact of government-imposed restrictions was more often highlighted in news stories of suicide than the virus itself (e.g. by causing reduced contact with others and disruption of routine). Employment and, to a lesser extent, financial concerns were also highlighted in such reports as having contributed to, or even caused, the suicidal behaviour. Concerns relating to education were relatively infrequent overall, but mentioned in a substantial proportion of news items focusing on young people under the age of 18 years (17/46, 40%). COVID-19 related bereavement, domestic conflict and housing-related problems were mentioned less frequently, as were fears about life post-COVID. No article focused on alcohol or drug-related issues. In almost half the articles, more than one COVID-19 related factor was reported to have played a role in suicidal behaviour (137/285, 48%: 86 (30.2%) mentioned two factors, 35 (12.3%) three, 10 (3.5%) four and 6 (2.1%) suggested five COVID-19 related influences).

In nearly two-thirds of news items a single ‘main issue’ from our list of potential COVID-19 related factors was suggested to have contributed to suicidal behaviour (181, 63.5%). Once again, lockdown-related restrictions were a more common theme than virus-specific fears (Table 2). Around one in ten stories reported frontline working, feelings of isolation, worsening mental health or employment-related difficulties (including job losses, furlough and redundancies) as key, potentially causative, factors in the suicidal behaviour. The impact of the pandemic on individuals’ education or finances was more rarely portrayed as a ‘main issue’. Interpersonal and domestic conflicts and housing-related problems were absent from the list of potential factors.

### 3.3 Quality of reporting

Over a quarter of articles included messages deemed to be ‘positive’ (79/285, 27.7%), particularly around encouraging help-seeking (n=33) and reaching out to loved ones (n=30), raising awareness of mental health and wellbeing (n=21) and related services/support (n=19), and around the importance of talking about such issues (n=16) and about suicide in particular (n=5). Based on comparison with international reporting guidelines, only a small number of articles were rated as ‘negative’ in relation to their headline (25/285, 8.8%), imagery (15/256, 5.9%) or overall coverage (8/285, 2.8%). However, examples of sensational language (69, 24.2%), over-simplistic explanations (93, 32.6%) and romanticised accounts of suicidal behaviour (7, 2.5%) were not uncommon. A substantial proportion of articles did not signpost readers to sources of help and support (117, 41.1%).

## 4. Discussion

We analysed British news coverage of possible COVID-19 related suicides and suicide attempts in the first four months of the pandemic after the week before lockdown in the UK. Our findings suggest that during this period approximately one in five news reports on suicides in the British press made a strong and direct link with the COVID-19 virus and associated restrictions, often in the headlines and imagery of such stories, as well as in the body of the articles themselves. The lockdown, with its inherent restrictions, was more frequently portrayed as a contributory or even causal factor in suicidal behaviour than the virus itself, particularly through the induction or exacerbation of feelings of isolation, entrapment and poor mental health. It is notable, however, that one in six articles focused on suicides involving healthcare staff, and one in ten highlighted the stress and trauma of working on the frontline at this challenging time.

Whilst a clear picture of the impact of the pandemic on specific occupational or socio-demographic groups is yet to emerge from official statistics and other scientific evidence, it is possible that news coverage is currently skewed towards frontline staff and, as also observed prior to COVID-19 (Marzano et al, 2018), towards young and relatively unusual suicides (including those involving a celebrity, murder, and violent methods). The relative paucity of reports focusing on bereaved individuals, women affected by domestic violence, financial, addiction and housing issues is perhaps surprising, but this may well change over time as the longer-term consequences of the pandemic develop.

Other aspects of reporting are also likely to change with time, not least as more inquests of possible-COVID-19 related suicides take place (potentially leading to more details of individual suicides emerging in the public domain), and as the immediate and longer-term impact of rising death tolls and public health restrictions unfold and reverberate.

Compared to earlier coverage of suicide in the UK (Marzano et al, 2018), news reports of possible COVID-19 related suicides in the first four months of the pandemic were less likely to include details of methods used for suicidal acts in the text (55.3% in 2012-13 versus 37% in the current analysis) or in headlines (20.6% in 2012-13 versus 9.1% in this study). They were also much more likely to signpost readers to help and support (7.5% in 2012-13 versus 58.9% in the current study). These are key recommendations in international media guidelines. The relative lack of method-related imagery in the reports is also a positive finding, as is the paucity of ‘photo galleries’ and textual links between different suicides during the early phases of the pandemic. This may reflect the fact that Samaritans provided specific COVID-19 and suicide briefings to the media early on in the pandemic (Samaritans 2020b). However, there remains room for improvement in these and other aspects of reporting, not least the omission of sensational language and over-simplistic explanations (observed in at least a quarter of possible COVID-19 related suicide reports) and the placing of suicide stories in less prominent positions within print media (also a well-established recommendation in relevant guidelines, but not the case in one in ten of the stories we analysed).

### 4.1 Strengths and Limitations

This study is the first systematic analysis of news coverage of COVID-19 related suicides in the UK at a time of heightened concern and speculation over the effects of the pandemic and associated restrictions on mental health (Brooks et al., 2020; Holmes et al., 2020), in the country with the highest overall death toll during the first wave of the pandemic relative to comparable countries (Raleigh, 2020). Our findings are based on a well-established, evidence-informed media monitoring database which captures all media reports of suicides and attempted suicides in the UK (Fraser et al., 2017). It may not, however, capture all important aspects of reporting, particularly in relation to the novel and rapidly evolving challenges precipitated by the pandemic and social distancing restrictions. Our understanding of what constitutes ‘positive’ and ‘negative’ news coverage of COVID-19 related suicidal behaviour may well require further refinement as time - and the pandemic - progress. Importantly, the perspectives of those most likely to be affected by such coverage should also be reflected in this process.

### 4.2 Future research

Ongoing research is needed to assess the quality and content of possible COVID-19 related suicide reporting over the longer-term, with feedback to those responsible where this deviates from recommended standards. More research is also needed to investigate the potential beneficial effects of media coverage of COVID-19 and suicide (Niederkrotenthaler et al., 2010), including the extent to which these may improve help-seeking at times of crisis, and how to maximise the benefits and minimise the harm of such reporting, and of suicide coverage more generally. Ideally, this should also extend to media campaigns and public messages given by members of the research community and health experts (including in public research outputs and publications) (Knipe et al., 2021), and in other countries and other forms of media (including fiction and social media).

### 4.3 Conclusions

Responsible reporting of suicide can save lives (Sisask and Värnik, 2012). This descriptive analysis of UK news coverage of possible COVID-19 related suicides in the first four months of the pandemic suggests that, although rarely of poor overall quality, current reporting may be biased towards young people and frontline staff. It also appears to be failing, to varying degrees, to follow media guidelines in relation to key, evidence-informed recommendations, including, for example, the omission of details of suicide methods and sensational language, and inclusion of potential sources of help and support information for readers. Whilst overall standards of reporting have improved over recent years (Fraser et al., 2017), and many reports examined in this study included messages deemed to be positive, the scale and reach of the current crisis underscores the urgent need for further improvements and more cautious reporting (Hawton et al, 2020).

Given the known risks of imitation and normalisation of suicidal behaviour through media reporting, and the potential for protective effects (Niederkrotenthaler et al., 2010), it is important to keep monitoring trends and biases in news and wider media portrayal of suicides throughout the pandemic and in its aftermath. The quality, as well as the content, of such coverage needs ongoing and systematic attention, and more research to better understand its impact on different audiences, with regular feedback to those responsible for producing media reports.

## Data Availability

For reasons of confidentiality, raw data are not available from the authors

## Contributors

All persons who meet authorship criteria are listed as authors, and all authors certify that they have participated sufficiently in the work to take public responsibility for the content, including participation in the concept, design, analysis, writing, or revision of the manuscript. In particular:

KH, LM & LF conceived the study and developed the basis for the protocol.

All authors reviewed and refined the study protocol and coding scheme.

MH, EH and YL coded the newspaper articles.

LM analysed the data and, with KH, wrote the initial version of the manuscript.

All authors contributed to the interpretation of results and revision of the article.

All authors approved the final article.

## Acknowledgments

The authors are very grateful to Jacqui Morrisey, Assistant Director, Research and Influencing at Samaritans, for her invaluable support and feedback, and Rachel Adamec, of the Samaritans Media Advisory Team, for all her help with coding newspaper articles.

